# Second versus first wave of COVID-19 deaths: shifts in age distribution and in nursing home fatalities

**DOI:** 10.1101/2020.11.28.20240366

**Authors:** John P.A. Ioannidis, Cathrine Axfors, Despina G. Contopoulos-Ioannidis

## Abstract

**OBJECTIVE:** To examine whether the age distribution of COVID-19 deaths and the share of deaths in nursing homes changed in the second versus the first pandemic wave.

**ELIGIBLE DATA:** We considered all countries that had at least 4000 COVID-19 deaths occurring as of January 14, 2020, at least 200 COVID-19 deaths occurring in each of the two epidemic wave periods; and which had sufficiently detailed information available on the age distribution of these deaths. We also considered countries with data available on COVID-19 deaths of nursing home residents for the two waves.

**MAIN OUTCOME MEASURES:** Change in the second wave versus the first wave in the proportion of COVID-19 deaths occurring in people <50 years (“young deaths”) among all COVID-19 deaths and among COVID-19 deaths in people <70 years old; and change in the proportion of COVID-19 deaths in nursing home residents among all COVID-19 deaths.

**RESULTS:** Data on age distribution were available for 14 eligible countries. Individuals <50 years old had small absolute difference in their share of the total COVID-19 deaths in the two waves across 13 high-income countries (absolute differences 0.0-0.4%). Their proportion was higher in Ukraine, but it decreased markedly in the second wave. The odds of young deaths was lower in the second versus the first wave (summary odds ratio 0.80, 95% CI 0.70-0.92) with large between-country heterogeneity. The odds of young deaths among deaths <70 years did not differ significantly across the two waves (summary odds ratio 0.95, 95% CI 0.85-1.07). Eligible data on nursing home COVID-19 deaths were available for 11 countries. The share of COVID-19 deaths that were accounted by nursing home residents decreased in the second wave significantly and substantially in 8 countries (odds ratio estimates: 0.22 to 0.66), remained the same in Denmark and Norway and markedly increased in Australia.

**CONCLUSIONS:** In the examined countries, age distribution of COVID-19 deaths has been fairly similar in the second versus the first wave, but the contribution of COVID-19 deaths in nursing home residents to total fatalities has decreased in most countries in the second wave.

## INTRODUCTION

Many countries around the world saw a pattern of the coronavirus disease 2019 (COVID-19) pandemic where a first wave occurred in the spring that substantially subsided during the summer, and a second wave emerged in the fall of 2020. A key question is whether the age distribution of COVID-19 fatalities in these locations remained steady between the two waves or not. COVID-19 has an extremely steep risk gradient for death across age groups (1-3). The relative share of infections among young, older, and debilitated people may shape the observed proportion of deaths in different demographic groups and the overall fatalities and infection fatality rate in the total population.

Data from a considerable number of seroprevalence studies done in different countries have suggested that not all age groups may have been equally infected during the spread of the virus in the first wave (4). In several countries that did very well in the first wave, e.g. Singapore, Australia, or Iceland (5) evidence suggests that elderly people were less likely to be infected and in particular nursing homes (long-term care facilities) were not contributing many fatalities (4). The opposite pattern was seen in countries that had high rates of death, where nursing homes were massively infected during the first wave (6). One wonders whether preferential protection of older, higher-risk people and in particular of nursing home residents is possible and whether this might have happened more efficiently in the second wave (7).

In order to answer these questions, we assessed the age distribution of COVID-19 fatalities in the two waves in countries with a substantial burden of fatalities as of January 2021, and we also assessed the relative contribution from deaths of nursing home residents in the two waves.

## METHODS

### Data for comparison of age distribution of COVID-19 deaths in second versus first wave

We considered data from publicly available situational reports of countries that had a large number of deaths in both the first and second waves, so as to allow meaningful inferences in comparing the age distributions, and where the trough between the two waves had happened between May 15 and September 15, 2020. Specifically, we considered all countries that had at least 4000 COVID-19 deaths, at least 200 COVID-19 deaths occurring in the first wave, and at least 200 COVID-19 deaths occurring in the second wave; and which had information available on the age distribution of these deaths separately. Searches were last updated on January 14, 2021. In order to separate the two wave periods in a consistent manner, we used the date between the two peaks that had the trough (lowest number of deaths) for a 7-day average according to Worldometer data (8). When two or more dates were tied for trough values, we picked the earliest one. In some countries, e.g. in the USA, for the second wave a separate peak may be discerned in late summer, followed by a higher peak in the late fall and winter. However, for consistency and convenience we separated all countries into two time periods called “first” and “second” wave.

For each eligible country, we found the most recent situational report that mentioned age distribution of deaths; and the situational report that mentioned age distribution of deaths as of the trough date (or a separation date for the two waves that was as close as possible to the trough, when data were not available specifically up to the trough date). It is acknowledged that a few deaths reported after the trough/separation dates may have happened earlier, but this is likely to pertain to very small numbers and it would not change the overall comparison.

Age cut-offs of interest were pre-specified to be 50 years and 70 years. In European countries that have seen two waves and in the USA, almost two-thirds of the population are younger than 50 years old and the age stratum of 50-69 accounts for another quarter of the population. In a few other countries that have also seen two waves, the proportion of the population younger than 50 years old even exceeds 80%, e.g. in Turkey or Pakistan. However, the lion’s share of COVID-19 deaths happen in people above 70; moreover, in countries where elder care facilities are common, a large share of COVID-19 deaths occur in nursing home residents.

Documentation of COVID-19 infection may be least systematic in this upper age stratum. The number of COVID-19 deaths may be most error prone and variable in its documentation across countries and even within the same country over time in elderly people, and nursing home residents in particular. Therefore, we aimed to assess whether (A) the proportion of COVID-19 deaths occurring in people <50 years old among all COVID-19 deaths changed in the second versus the first wave; and (B) whether the proportion of COVID-19 deaths occurring in people <50 years old among the COVID-19 deaths occurring in people <70 years old changed between the two waves. When data were not provided for the cut-offs of 50 and 70 years for COVID-19 deaths, we used the closest cut-offs available provided it was not more than 5 years off (45 and 65 years).

### Data for COVID-19 deaths in nursing home residents in second versus first wave

We also extracted data on COVID-19 deaths occurring in nursing home residents. We preferred data on all COVID-19 deaths of such residents (occurring in the nursing homes or in hospitals) unless information was available only for COVID-19 deaths happening in nursing homes. We also preferred data on both confirmed and probable COVID-19 deaths, unless only data on confirmed deaths were available.

We compared proportions of these nursing home residents’ deaths among all COVID-19 deaths in the second versus first wave periods. The International Long-Term Care Policy Network has issued reports based on available country-level official data for the number of deaths among nursing home residents linked to COVID-19. Their previous report (June 26, 2020) (9) was considered. Given the substantial spread of COVID-19 occurring in many countries after their latest report, we restricted our analysis to the countries for which we could find updates that covered at least until the end of 2020 using the official sources cited in the International Long-Term Care Policy Network reports. Latest extraction of these data was performed between January 14 and January 20, 2021. We used as cutoff for the first wave the dates given in the June 26 report (between June 1 and June 23, 2020).

Of note, we use here the term “nursing home” broadly to include different types of long-term care facilities. The exact types of facilities included in the COVID-19 death data for each country may differ and we recorded the definition used in each country. We ensured also that the definition remained the same in the two waves, and recorded any noted changes in the documentation of COVID-19 deaths in nursing homes.

### Statistical analyses

All comparisons of proportions of COVID-19 deaths per country between the two periods used odds ratios and 95% confidence intervals thereof. Meta-analysis of odds ratios used a random effects model. Heterogeneity was expressed with the I^2^ statistic and tested with the chi-squared-based Q test. P-values are two-tailed. Statistical analyses were run in STATA (10).

## RESULTS

### Eligible data for age distribution of COVID-19 deaths in the two waves

49 countries had at least 4000 deaths until January 14, 2021. Of those, 22 countries (Austria, Belgium, Canada, Czechia, France, Germany, Hungary, Italy, Japan, Netherlands, Pakistan, Poland, Portugal, Romania, Russia, Spain, Sweden, Switzerland, Turkey, Ukraine, United Kingdom, USA) also had at least 200 deaths in each wave and had a trough between May 15 and September 15. We could retrieve some data on the age distribution separately for the two waves for 17 of the 22 countries. However, we could not retrieve reports for Canada for deaths at age <50 in the first wave; Romania provided only graphs with percentages per age group and resolution was not sufficiently accurate in the <50 years old age group; and for Turkey there were no age-stratified data available after late October. Eventually, data from 14 countries were finally included (Table 1). As shown in Table 1, the trough separating the two waves occurred between June 1 and August 28 in all locations. The number of deaths in the second wave as of the date of the analysis was higher than the number of deaths in the first wave in all countries with eligible data, except for Spain and Sweden. Sources of information for the 14 countries appear in Supplementary Table 1.

**Table 1.** Eligible locations for analyses of COVID-19 deaths per age group

| Countries | Date of<br>trough | Deaths<br>on<br>trough | Deaths<br>in first<br>wave | Deaths in<br>second<br>wave | Separation<br>date* | Latest<br>date** |
| --- | --- | --- | --- | --- | --- | --- |
| Austria | June 5 | 0 | 645 | 6183 | June 5 | Jan 14 |
| Belgium | July 14 | 1 | 9671 | 10623 | July 14 | Jan 14 |
| France | Aug 10 | 6 | 30354 | 38448 | Aug 11 | Jan 12 |
| Germany | Aug 1 | 3 | 9148 | 32429 | Aug 1 | Jan 12 |
| Italy | Aug 15 | 5 | 35614 | 39434 | Aug 30 | Jan 5 |
| Japan | July 6 | 0 | 977 | 2742 | July 6 | Jan 6 |
| Netherlands | July 14 | 0 | 6135 | 6428 | May 22 | Jan 12 |
| Portugal | Aug 7 | 1 | 1750 | 6634 | Aug 7 | Jan 13 |
| Spain | July 27 | 1 | 29856 | 23025 | July 27 | Jan 13 |
| Sweden | Aug 28 | 1 | 5739 | 4364 | Aug 28 | Jan 14 |
| Switzerland | June 1 | 0 | 1831 | 6755 | May 23 | Jan 14 |
| Ukraine | June 3 | 11 | 735 | 16437 | June 3 | Dec 23 |
| United Kingdom | Aug 21 | 7 | 56653 | 32590 | Aug 21 | Jan 1 |
| USA | July 5 | 518 | 130951 | 198660 | July 4 | Jan 9 |

\*Data were not always available to separate deaths in the first versus the second wave using the trough date,
and in these cases the most proximal date with data available was used. \*\*Data were not always available until
January 14, and in these cases the most proximal date with available data was used.

### Age distribution of COVID-19 deaths in the two waves

Table 2 shows the age distribution of COVID-19 deaths in the first and second wave in each eligible country with data.

**Table 2.**
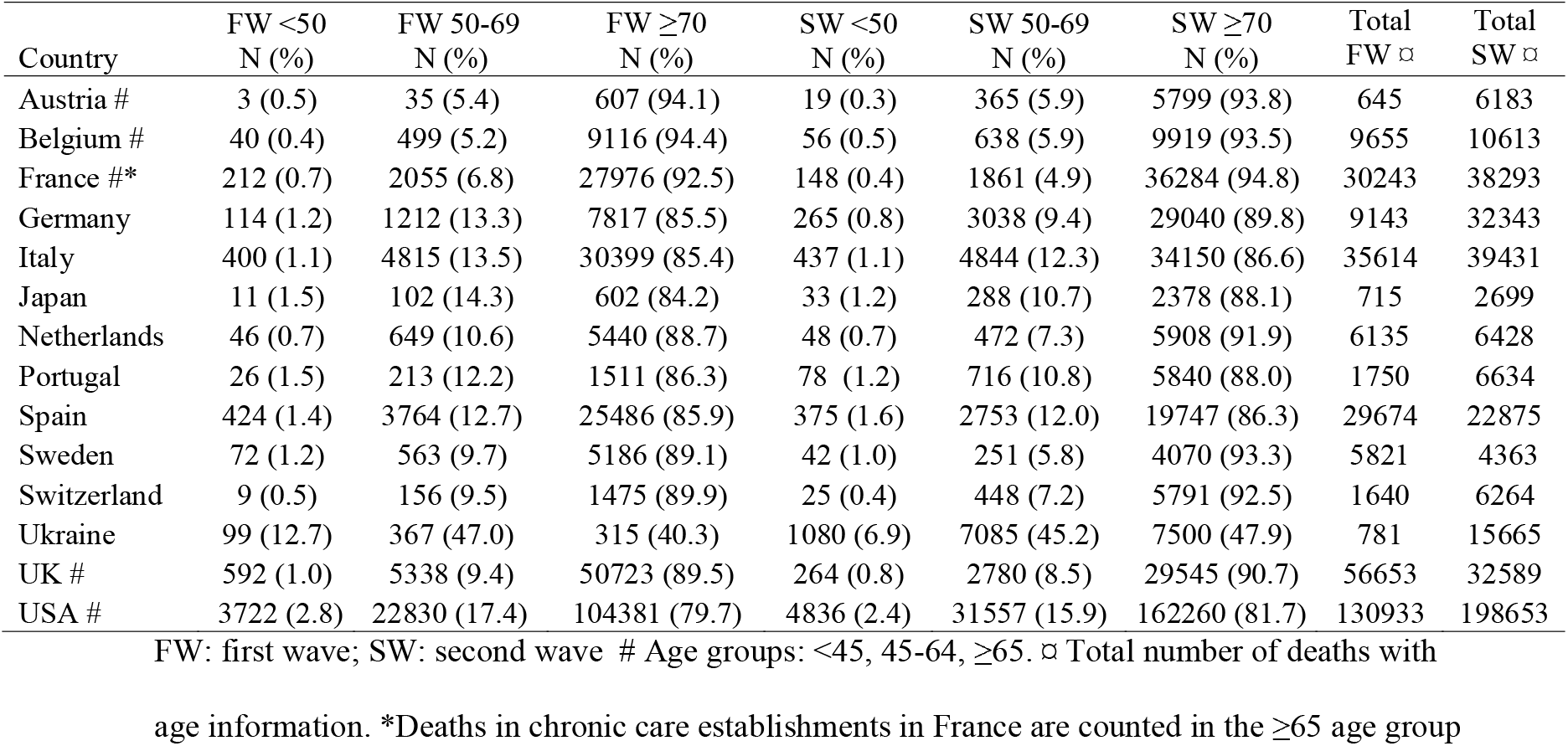
Proportion of COVID-19 deaths in specific age groups

The proportion of COVID-19 deaths <50 years among all COVID-19 deaths in the first wave did not exceed 1.5% in any high-income country except for the USA (2.8% for a cut-off of <45 years) and it was much higher in Ukraine (12.7%). In the second wave, the absolute difference versus the first wave was only 0.0-0.4% in high-income countries, with the largest decreases in the share of deaths <50 years occurring in USA (2.4% from 2.8% in the first wave) and in Germany (0.8% from 1.2%). A far more major decrease was seen in Ukraine (from 12.7% to 6.9%). As shown in Figure 1a, there was very large heterogeneity when results were expressed in an odds ratio comparing the two waves (I^2^=82%, p<0.001 for heterogeneity) and the summary odds ratio was 0.80, 95% CI 0.70-0.92, suggesting fewer young deaths in the second wave. In 5 countries (Ukraine, France, Germany, UK, USA) the odds were significantly less for young deaths in the second wave, while in Spain there was a borderline significance in the opposite direction.

**Figure 1.**
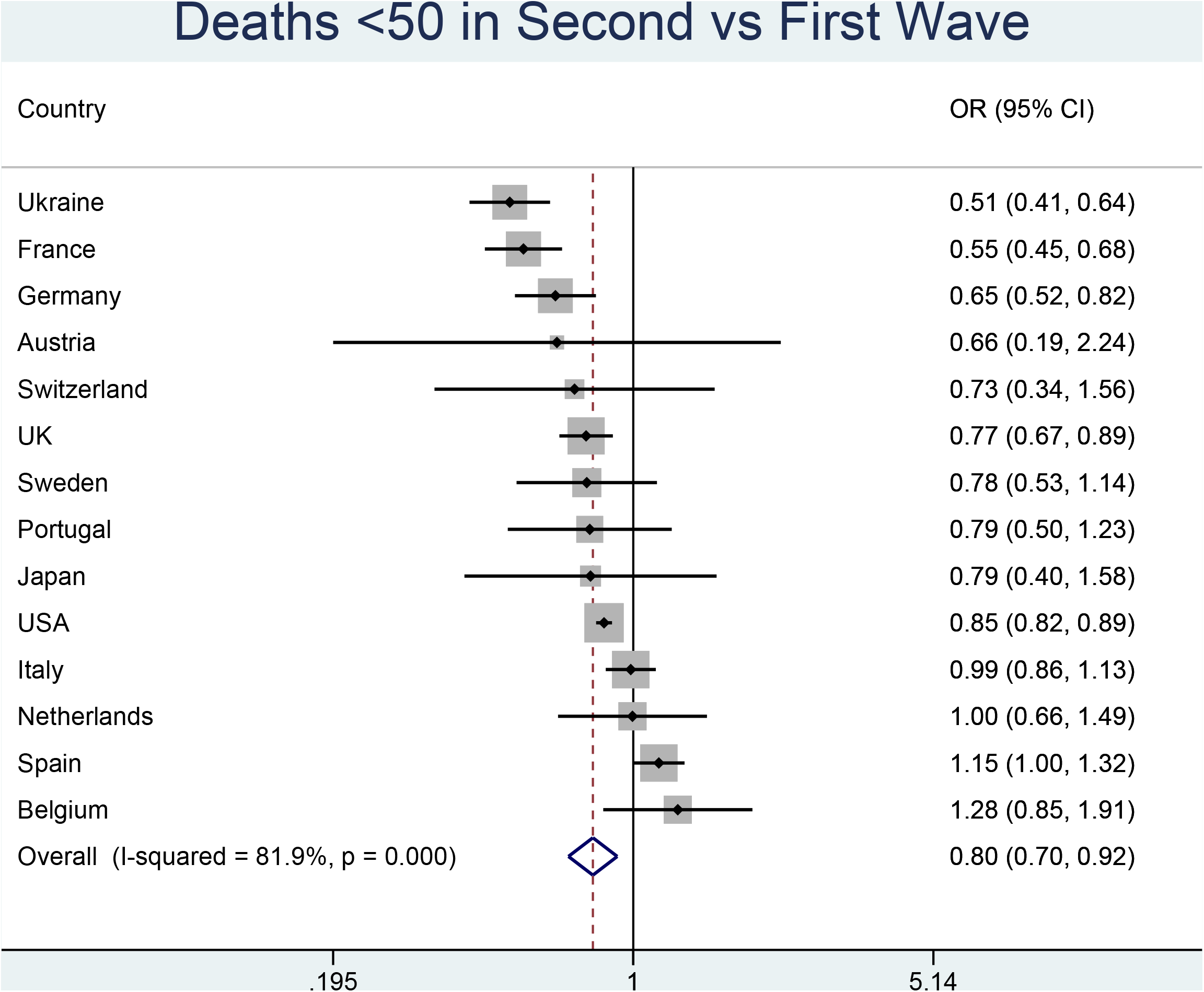
Panel a: COVID-19 deaths in people <50 years in the second versus first wave. Panel b: COVID-19 deaths in people <50 years among deaths in people <70 years in the second versus first wave.

When estimated only among the COVID-19 deaths in people <70 years old, the odds of COVID-19 deaths in individuals <50 years old showed no major differences on average in the second wave versus the first wave (Figure 1b) with a summary estimate of the odds ratio of 0.95 (95% CI 0.85-1.07). There was again significant between-country heterogeneity (I^2^=72%, p<0.001 for heterogeneity). In 4 countries (Ukraine, France, UK, USA) the odds were significantly less for young deaths in the second wave, while the opposite pattern was seen in Spain.

### COVID-19 deaths of nursing homes residents

Out of 21 countries with any documented COVID-19 nursing home deaths as per the International Long-Term Care Policy Network report on June 26 (9), eligible data on nursing home residents’ COVID-19 deaths in the first versus second wave could be obtained for 11 countries (Table 3). Sources of data appear in Supplementary Table 2. The definitions of COVID-19 deaths and nursing home institutions differed across the different countries. In four countries, nursing home deaths data included only deaths that occurred in the nursing home environment (i.e. in-hospital deaths of nursing home residents were excluded). In five countries, only confirmed COVID-19 nursing home deaths were considered. Definitions of nursing home facilities and other included facilities appear also in Table 3.

**Table 3.**
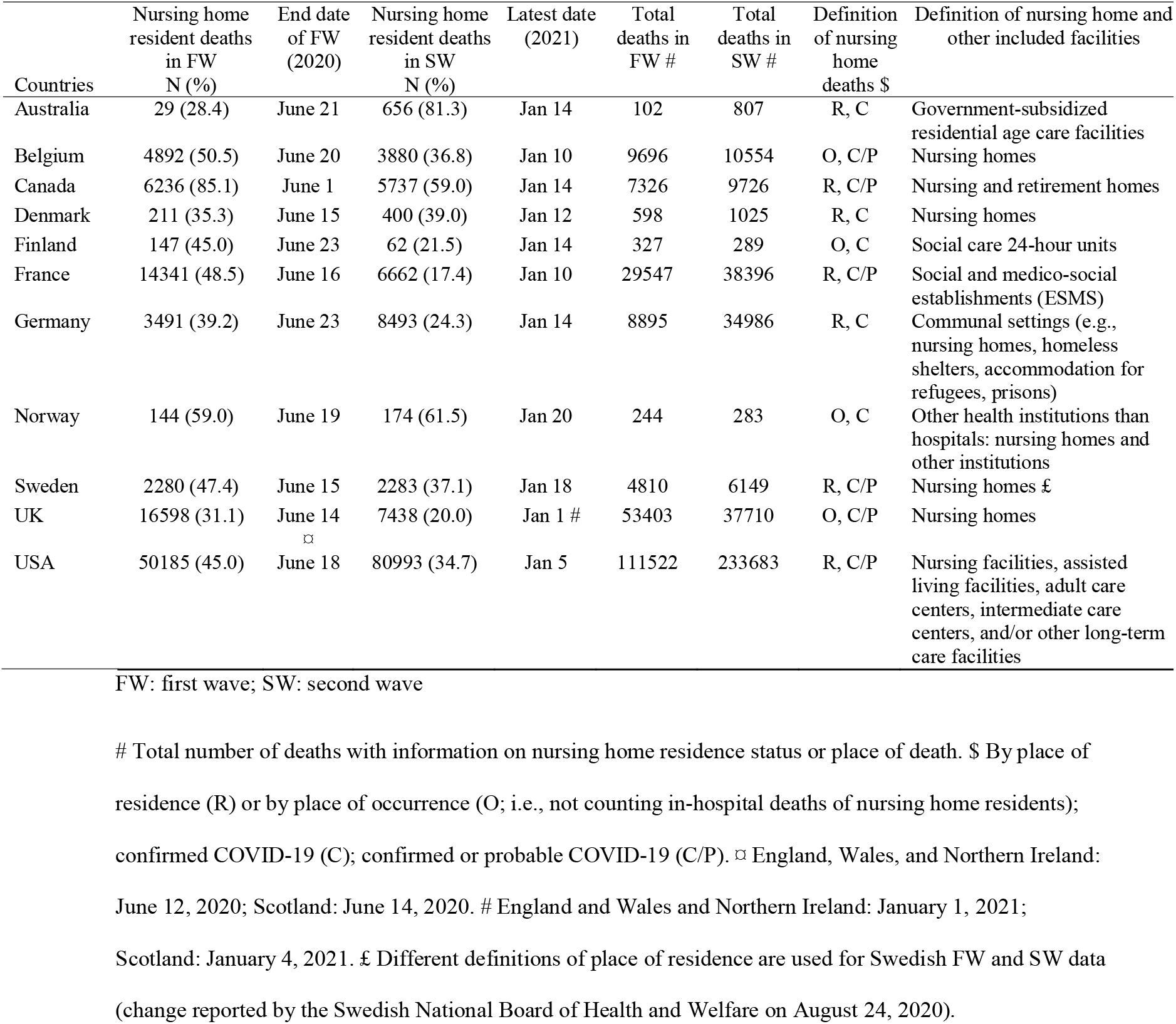
Proportion of COVID-19 deaths occurring in nursing home residents

| Countries | Nursing home resident deaths in FW N (%) | End date of FW (2020) | Nursing home resident deaths in SW N (%) | Latest date (2021) | Total deaths in FW # | Total deaths in SW # | Definition of nursing home deaths \$ | Definition of nursing home and other included facilities |
| --- | --- | --- | --- | --- | --- | --- | --- | --- |
| Australia | 29 (28.4) | June 21 | 656 (81.3) | Jan 14 | 102 | 807 | R, C | Government-subsidized residential age care facilities |
| Belgium | 4892 (50.5) | June 20 | 3880 (36.8) | Jan 10 | 9696 | 10554 | O, C/P | Nursing homes |
| Canada | 6236 (85.1) | June 1 | 5737 (59.0) | Jan 14 | 7326 | 9726 | R, C/P | Nursing and retirement homes |
| Denmark | 211 (35.3) | June 15 | 400 (39.0) | Jan 12 | 598 | 1025 | R, C | Nursing homes |
| Finland | 147 (45.0) | June 23 | 62 (21.5) | Jan 14 | 327 | 289 | O, C | Social care 24-hour units |
| France | 14341 (48.5) | June 16 | 6662 (17.4) | Jan 10 | 29547 | 38396 | R, C/P | Social and medico-social establishments (ESMS) |
| Germany | 3491 (39.2) | June 23 | 8493 (24.3) | Jan 14 | 8895 | 34986 | R, C | Communal settings (e.g., nursing homes, homeless shelters, accommodation for refugees, prisons) |
| Norway | 144 (59.0) | June 19 | 174 (61.5) | Jan 20 | 244 | 283 | O, C | Other health institutions than hospitals: nursing homes and other institutions |
| Sweden | 2280 (47.4) | June 15 | 2283 (37.1) | Jan 18 | 4810 | 6149 | R, C/P | Nursing homes £ |
| UK | 16598 (31.1) | June 14<br>⌘ | 7438 (20.0) | Jan 1 # | 53403 | 37710 | O, C/P | Nursing homes |
| USA | 50185 (45.0) | June 18 | 80993 (34.7) | Jan 5 | 111522 | 233683 | R, C/P | Nursing facilities, assisted living facilities, adult care centers, intermediate care centers, and/or other long-term care facilities |

FW: first wave; SW: second wave

The proportion of nursing home COVID-19 deaths among all COVID-19 deaths was lower in the second wave than in the first wave in 8 of the 11 countries (Figure 2). There were large and statistically significant odds reductions in these 8 countries (odds ratios 0.22 to 0.66). There were no significant differences in Denmark and Norway where few deaths overall were recorded in both waves, and a major increase in the proportion of nursing home COVID-19 deaths in Australia (odds ratio 10.9, 95% CI 6.9-17.4) in the second wave. This resulted in extreme between-country heterogeneity (I^2^=99.7%, p<0.001 for heterogeneity) making a single summary odds ratio not meaningful to obtain.

**Figure 2.**
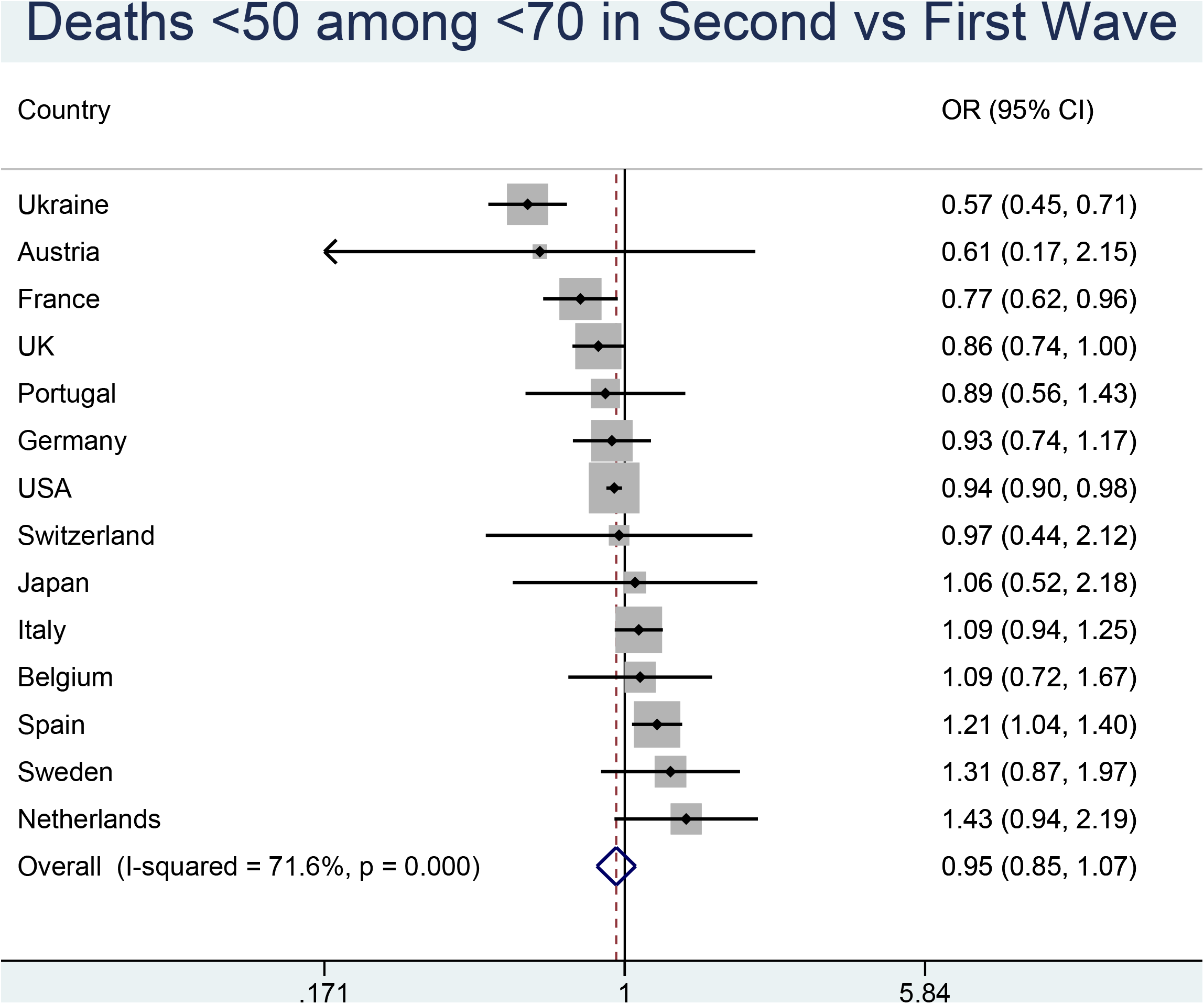
COVID-19 deaths in nursing home residents in the second versus first wave.

## DISCUSSION

### Main findings

The age distribution of COVID-19 deaths did not change much within the examined high-income countries between the first and second waves. Deaths in individuals <50 years accounted for approximately 1% of all deaths in European high-income countries, and modestly higher proportion in the USA. Some countries even saw modest decreases of this percentage in the second wave compared with the first. Concurrently, there was a strong pattern for a decreasing share of COVID-19 deaths of nursing home residents in the second wave versus the first wave, with Australia being an exception to this pattern. Data from one middle-income country (Ukraine) showed overall much higher proportion of COVID-19 deaths among people <50 years old, with a substantial decrease in the second wave.

### Nursing home COVID-19 deaths

The decreasing share of nursing home resident COVID-19 deaths in the second wave may reflect multiple factors. Higher awareness of the extreme fatality risk of nursing home residents and raised efforts to protect nursing homes (learning from experience) (11, 12) may be primarily responsible. Perhaps hygiene measures, infection control, testing of personnel and residents, and avoidance of staff working across multiple nursing homes have become more routine during the second wave. Another potential contributing factor may be better treatment and management. For example, dexamethasone (13) has been available in the second wave; conversely, use of mortality-increasing hydroxychloroquine (14) and suboptimal mechanical ventilation practices may have been reduced in some countries. Cohort effects are also possible: e.g. the first wave may have killed some of the most frail residents and/or may have already preferentially devastated nursing homes with poorer standards and thus higher infection risk. In countries with the heaviest toll of nursing home deaths, a large share of nursing home residents and personnel (perhaps even the majority) may have been infected by January 2021. E.g. according to the COVID Tracking Project (15) as of January 21, there are 1,183,661 documented COVID-19 cases and 146,294 COVID-19 deaths in nursing homes in the USA. With such extremely high infection rates, the residual pool of infected nursing home residents has shrunk markedly, and this may explain in part the decreasing share of nursing home deaths to total COVID-19 deaths.

Some additional factors need to be considered. Within the same country, the second wave may have spread to different communities with different shares of nursing home populations compared with the first wave. For example, in the USA there have been distinct waves of the pandemic that have impacted various regions, e.g., northeast and northwest in the spring of 2020, the sunbelt region in the summer, the Midwest and California in the late summer, and widespread outbreaks by late fall/winter 2020, especially in areas that had been relatively spared earlier. Also some data artefact cannot be fully excluded, in particular if some nursing home COVID-19 deaths in the second wave suffer reporting delays. Finally, we tried to ensure that definitions of deaths and nursing home facilities were similar in the two waves, but subtle changes cannot be excluded, e.g. Sweden made some changes in the defining documentation of nursing home deaths during the summer months. Regardless, the difference between the two waves in overall COVID-19 deaths in most countries is quite large, thus unlikely to be only a data or methods artefact.

Despite the clear improvement, the proportion of COVID-19 deaths that occurred among nursing home residents remained very large in western European countries and the USA. Moreover, not all countries have seen improvements in the second wave. Australia witnessed a major increase, and this may be explained by non-remedied dysfunctions in its elder care (16). The Australian aged care system has long been criticized for understaffing, and using low-pay staff with poor skills who work across multiple facilities. Most facilities are privately run for-profit enterprises. Similar problems with nursing home care inefficiencies have been described also in other countries with a large share of deaths in nursing homes, e.g. in Canada (17); however, in other countries, COVID-19 already hit nursing homes rapidly during the first wave.

### Age distribution

The relative stability of the age distribution of COVID-19 deaths in the same country between the two waves suggests that country-specific population demographics are the key driver of the age distribution of infections, which then get reflected also in the age distribution of deaths (18). Other features that may also affect COVID-19 mortality and its age distribution (e.g. population density, deprivation, ethnicity, frequencies of pre-existing conditions, occupational profile, and environmental factors) also remain steady within the same country between the two waves.

While the absolute share of deaths at age <50 among all COVID-19 deaths had small fluctuations between the two waves, in relative terms several countries (Germany, USA, UK, France, Ukraine) had a smaller share of young deaths in the second wave. One may expect shifts in the age distribution of COVID-19 deaths in the second wave if the epidemic within a country is spreading to new epicenters and populations with different demographics. Alternatively, documentation of COVID-19 may have become more aggressive in the second wave with more testing being performed, especially among deceased elderly.

We had data only from one middle-income country (Ukraine) and no data from low-income countries. Differential demographics in the two waves (e.g. higher infection rates in urban and congested young populations in the first wave, followed by older provincial populations later) may be more prominent in lower-income countries. For example, Turkey is another country with very high proportion of deaths occurring in young people in the first wave (6.8% among those <50). During the first wave, the highest numbers of cases per million population were seen in Istanbul and southern Turkey (19). Both epicenters are characterized by a very young population, younger than the inhabitants of other locations that were hit more in the second wave. Data on age distribution of deaths were available for Turkey only until late October, showing a decrease of the proportion of deaths among those <50 years (4.8%). Alternatively, non-documentation of COVID-19 deaths, especially among the elderly and even more so in the first wave when testing was more limited might have been even more prominent in developing than in high-income countries.

### Implications

The observed patterns of diminished COVID-19 deaths in nursing homes in many countries may act in the direction of decreasing the infection fatality rate of the pandemic in several high-income countries in the second wave and any subsequent waves that might occur. For an equal number of total infections, when nursing homes are spared, the number of total COVID-19 deaths will be substantially less (20). With prioritization of vaccination for nursing home residents and elderly individuals in early 2021, the share of nursing home deaths and of deaths among the elderly in general may decrease further. This may induce an increase in the share of young deaths among the total. Paradoxically, having a larger share of young deaths would not be a bad sign, but may indirectly be an indicator of better protection of vulnerable elderly individuals. This pattern will not be observed, however, if uptake of vaccines is not preferentially higher among the most vulnerable, if vaccines are less effective among the most vulnerable, and if there is larger risk compensation among the vulnerable who are vaccinated (e.g. marked decrease in other protections taken with non-pharmaceutical measures). For countries where the pandemic shifts from urban centers with young population to areas with older populations, infection fatality rate may increase. The same applies to any countries that successfully protected their nursing homes in the first wave, but have been less successful in the second wave.

Some measures taken to contain the pandemic may affect differentially people at different ages and may have different real-world effectiveness for people at different ages. More data need to be accumulated as the pandemic progresses, as different types of lockdown measures are used and relieved, and as vaccines become more widely deployed. Some countries like Italy have presented data where the median age of fatalities during the first wave started at lower values, increased during draconian lockdown, and then declined again as restrictions were relieved (21). Such differences over time may have been due to chance. However, an alternative explanation might be that draconian lockdown increases the level of protection more prominently for the young than for the elderly compared with pre- or post-lockdown behavior. E.g. the elderly may be taking severe precautions anyhow, regardless of government-imposed lockdown status.

### Limitations

Some limitations need to be discussed. First, age information was missing on some deaths, but this pertained to very few fatalities and it is unlikely to have created systematic bias in the comparison of first versus second waves. Second, many nursing home deaths have substantial ambiguity in their attribution to COVID-19, especially when there is no test confirmation. We tried to use data with consistent definitions and approaches in the two waves, but the two periods may still differ, e.g. typically more testing was done in the second wave. If anything, this would usually tend to increase the number of confirmed COVID-19 deaths in nursing homes in the second wave. Third, it would be useful to understand whether there are differences between the two waves in the share of deaths in people without underlying conditions and/or specific risk profiles. This could give additional insights about the relative exposure and protection of these groups in the two time periods. However, such data are very sparse in currently available situational reports (e.g., 22, 23). Fourth, we did not find sufficiently detailed data on age distribution of COVID-19 deaths even for some high-income countries. This is a deficiency that could be quickly corrected in country-level situational reports worldwide.

Finally, we found very sparse data from middle-income countries and no data from low-income countries. However, most low-income countries have not had a trough separating two waves. Instead, they have typically seen continuous epidemic activity comprising a single wave. The share of COVID-19 death that are accounted by young people is probably greater in middle- and low-income countries and in countries with many impoverished people. E.g., in Colombia (an upper middle-income country), 8% of COVID-19 deaths are accounted by ages <50 years and the proportion is 6% even in Chile which qualifies for a high-income country but has a large rich-poor gap and many impoverished citizens (24, 25); in Indonesia (a country transitioning from lower to upper middle-income country), 20.4% of COVID-19 deaths are accounted by ages <45 years (26); and in India (a lower middle-income country), 14.4% of COVID-19 deaths are accounted by ages <40 (2). Moreover, in all countries disadvantaged people with lower socioeconomic status are often more hit by the pandemic (as well as the measures taken, e.g. in terms of unemployment, loss of health insurance, food insecurity, mental health burden, housing instability, and loss of preventive healthcare). Overall, during 2020 only ∼1% of COVID-19 deaths in high-income European countries were in ages <50 (accounting for only ∼4,000 fatalities), while the overall proportion across the rest of the world may have been ∼5% or higher (accounting for ∼100,000 fatalities). The impact of measures taken on mid-term and long-term mortality worldwide needs careful study and it may affect more prominently young people than COVID-19 itself.

### Future questions

Acknowledging these caveats, demographic profile changes in the future evolution of the COVID-19 pandemic activity should be monitored. Moreover, the effects of different non-pharmaceutical measures, as well as prospective vaccinations during 2021, on the distribution of deaths warrants close attention. It would be useful to understand the extent to which the demographic footprint of fatalities can be efficiently modulated by appropriate interventions in different settings.

## Data Availability

All the data are in the manuscript and tables.

## CRediT authorship contribution statement

John P.A. Ioannidis: Conceptualization, Methodology, Formal analysis, Investigation, Writing - original draft, Project administration.

Cathrine Axfors: Conceptualization, Software, Formal analysis, Investigation, Data curation, Writing - review & editing.

Despina G. Contopoulos-Ioannidis: Conceptualization, Methodology, Investigation, Data curation, Writing - review & editing.

---

**Supplementary Table 1.** Sources for COVID-19 deaths in specific age groups

| Country, first wave (FW) and/or second wave (SW) | Reference |
| --- | --- |
| Austria, FW and SW | Austria_2021_01_14_Deaths_Covid_19.xlsx [Dataset version 2021-01-14] Retrieved from https://dc-covid.site.ined.fr/en/data/austria |
| Belgium, FW | COVID19BE_MORT.csv [Dataset version 2020-11-14] Retrieved from: https://epistat.wiv-isp.be/covid/ |
| Belgium, SW | COVID19BE_MORT.csv [Dataset version 2021-01-14] Retrieved from: https://epistat.wiv-isp.be/covid/ |
| France, FW | Santé publique France. Point épidémiologique hebdomadaire du 13 août 2020. Retrieved from: https://www.santepubliquefrance.fr/recherche/#search=COVID%2019%20%20point%20epidemiologique&publications=donn%C3%A9es&regions=National&sort=date |
| France, SW | Santé publique France. Point épidémiologique hebdomadaire du 14 janvier 2021. Retrieved from: https://www.santepubliquefrance.fr/recherche/#search=COVID%2019%20%20point%20epidemiologique&publications=donn%C3%A9es&regions=National&sort=date |
| Germany, FW | Robert Koch Institut. Täglicher Lagebericht des RKI zur Coronavirus-Krankheit-2019 (COVID-19). 01.08.2020 – AKTUALISIERTER STAND FÜR DEUTSCHLAND |
| Germany, SW | Robert Koch Institut. Data obtained via The Demography of COVID-19 Deaths. National Institute for Demographic Studies (INED) (distributor). Extract from: https://dc-covid.site.ined.fr/en/ [2021-01-23] |
| Italy, FW | Istituto Superiore di Sanità (ISS). Epidemia COVID-19. Aggiornamento nazionale, 20 maggio 2020 – ore 16:00. |
| Italy, SW | Istituto Superiore di Sanità (ISS). Aggiornamento nazionale, 5 gennaio 2021 – ore 12:00. Retrieved from: https://www.epicentro.iss.it/en/coronavirus/sars-cov-2-integrated-surveillance-data |
| Japan, FW | Japan FW_jpn_death_share.csv [Dataset version 2020-08-18] Retrieved from: https://github.com/fumiyau/COVerAGE-JP. Reference: Uchikoshi 2020. “COVerAGE-JP: COVID-19 Deaths by Age and Sex in Japan” (SocArXiv) DOI: 10.31235/osf.io/cpqrt |
| Japan, SW | demography.csv [Dataset version 2021-01-14] Retrieved from: https://github.com/kaz-ogiwara/covid19/blob/master/data/demography.csv |
| Netherlands, FW | Rijksinstituut voor Volksgezondheid en Milieu – RIVM. Epidemiologische situatie COVID-19 in Nederland, 14 juli 2020, 10:00. Retrieved from: https://www.rivm.nl/coronavirus-covid-19/actueel/wekelijkse-update-epidemiologische-situatie-covid-19-in-nederland |
| Netherlands, SW | Rijksinstituut voor Volksgezondheid en Milieu – RIVM. Epidemiologische situatie van SARS-CoV-2 in Nederland. 12 januari 2021, 10:00. Retrieved from: https://www.rivm.nl/coronavirus-covid-19/actueel/wekelijkse-update-epidemiologische-situatie-covid-19-in-nederland |
| Portugal, FW and SW | Serviço Nacional de Saúde/Direção-Geral da Saúde. Data obtained via The Demography of COVID-19 Deaths. National Institute for Demographic Studies (INED) (distributor). Extract from: https://dc-covid.site.ined.fr/en/ [2021-01-23] |
| Spain, FW and SW | Ministerio de Sanidad / Instituto de Salud Carlos III. Data obtained via The Demography of COVID-19 Deaths. National Institute for Demographic Studies (INED) (distributor). Extract from: https://dc-covid.site.ined.fr/en/ [2021-01-23] |
| Sweden, FW and SW | Folkhälsomyndigheten / Socialstyrelsen. Data obtained via The Demography of COVID-19 Deaths. National Institute for Demographic Studies (INED) (distributor). Extract from: https://dc-covid.site.ined.fr/en/ [2021-01-23] |
| Switzerland, FW | 200325_Datengrundlage_Grafiken_COVID-19-Bericht.xlsx [Dataset version 2020-05-23] Retrieved from: https://www.bag.admin.ch/bag/de/home/krankheiten/ausbrueche-epidemien-pandemien/aktuelle-ausbrueche-epidemien/novel-cov/situation-schweiz-und-international.html#-1222424946 |
| Switzerland, SW | 200325_Datengrundlage_Grafiken_COVID-19-Bericht.xlsx [Dataset version 2021-01-14] Retrieved from: https://www.bag.admin.ch/bag/de/home/krankheiten/ausbrueche-epidemien-pandemien/aktuelle-ausbrueche-epidemien/novel-cov/situation-schweiz-und-international.html#-1222424946 |
| Ukraine, FW | Deaths-Age-Sex_Covid-19_Ukraine_17-11.xlsx [Dataset version 2020-11-25] Retrieved from: https://dc-covid.site.ined.fr/en/data/ukraine/ |
| Ukraine, SW | Deaths-Age-Sex_Covid-19_Ukraine_23_12.xlsx [Dataset version 2020-12-23] Retrieved from: https://dc-covid.site.ined.fr/en/data/ukraine/ |
| United Kingdom, FW | publishedweek432020.xlsx [Dataset version 2020-11-13] Retrieved from: https://www.ons.gov.uk/peoplepopulationandcommunity/birthsdeathsandmarriages/deaths/datasets/weeklyprovisionalfiguresondeathsregisteredinenglandandwales |
| United Kingdom, SW | publishedweek532020.xlsx [Dataset version 2021-01-14] Retrieved from: https://www.ons.gov.uk/peoplepopulationandcommunity/birthsdeathsandmarriages/deaths/datasets/weeklyprovisionalfiguresondeathsregisteredinenglandandwales |
| USA, FW and SW | Provisional_COVID-19_Death_Counts_by_Sex__Age__and_Week.csv [Dataset version 2021-01-14] Retrieved from: https://data.cdc.gov/NCHS/Provisional-COVID-19-Death-Counts-by-Sex-Age-and-W/vsak-wrfu |

**Supplementary Table 2.**
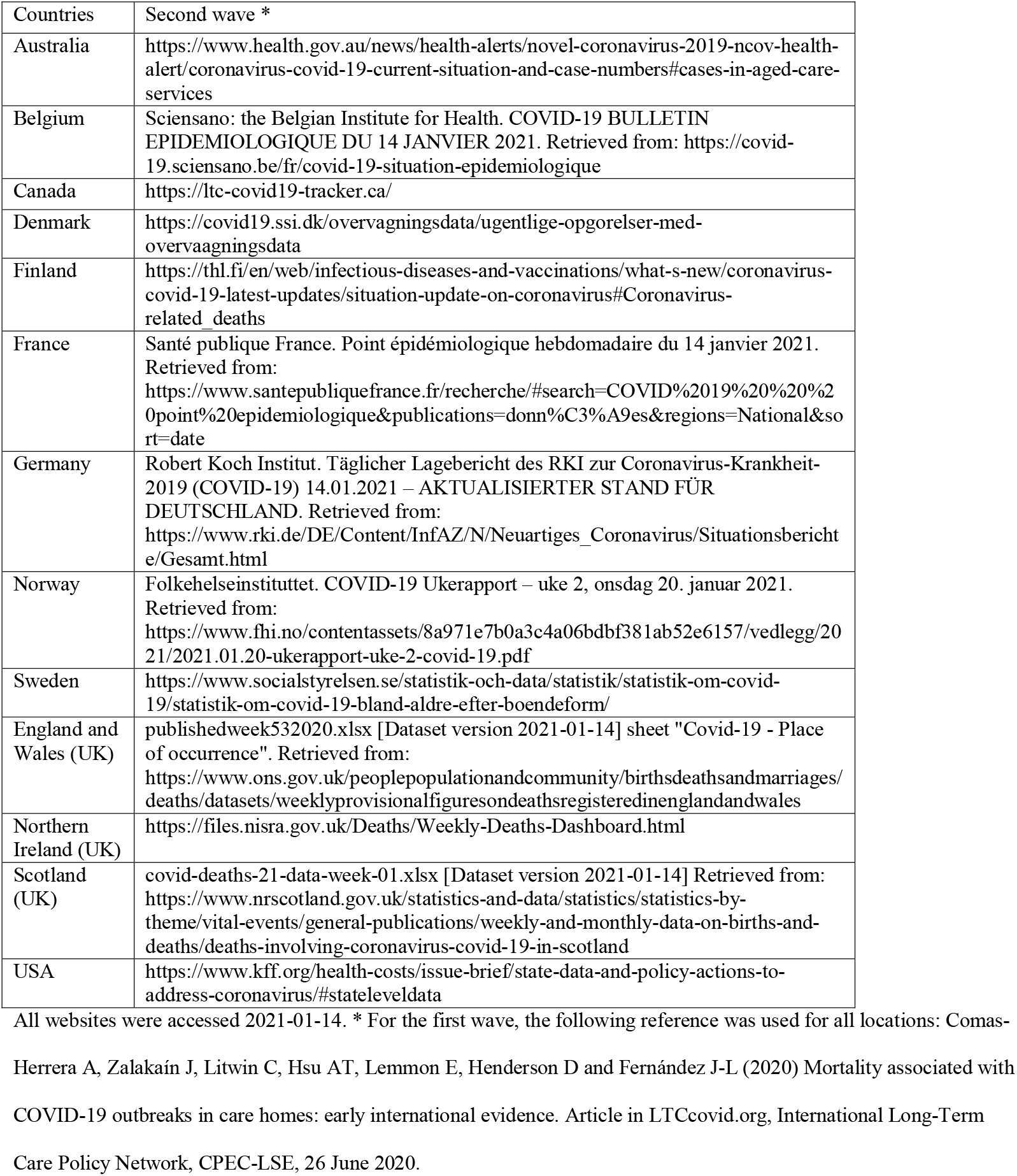
Sources for COVID-19 deaths occurring in nursing home residents

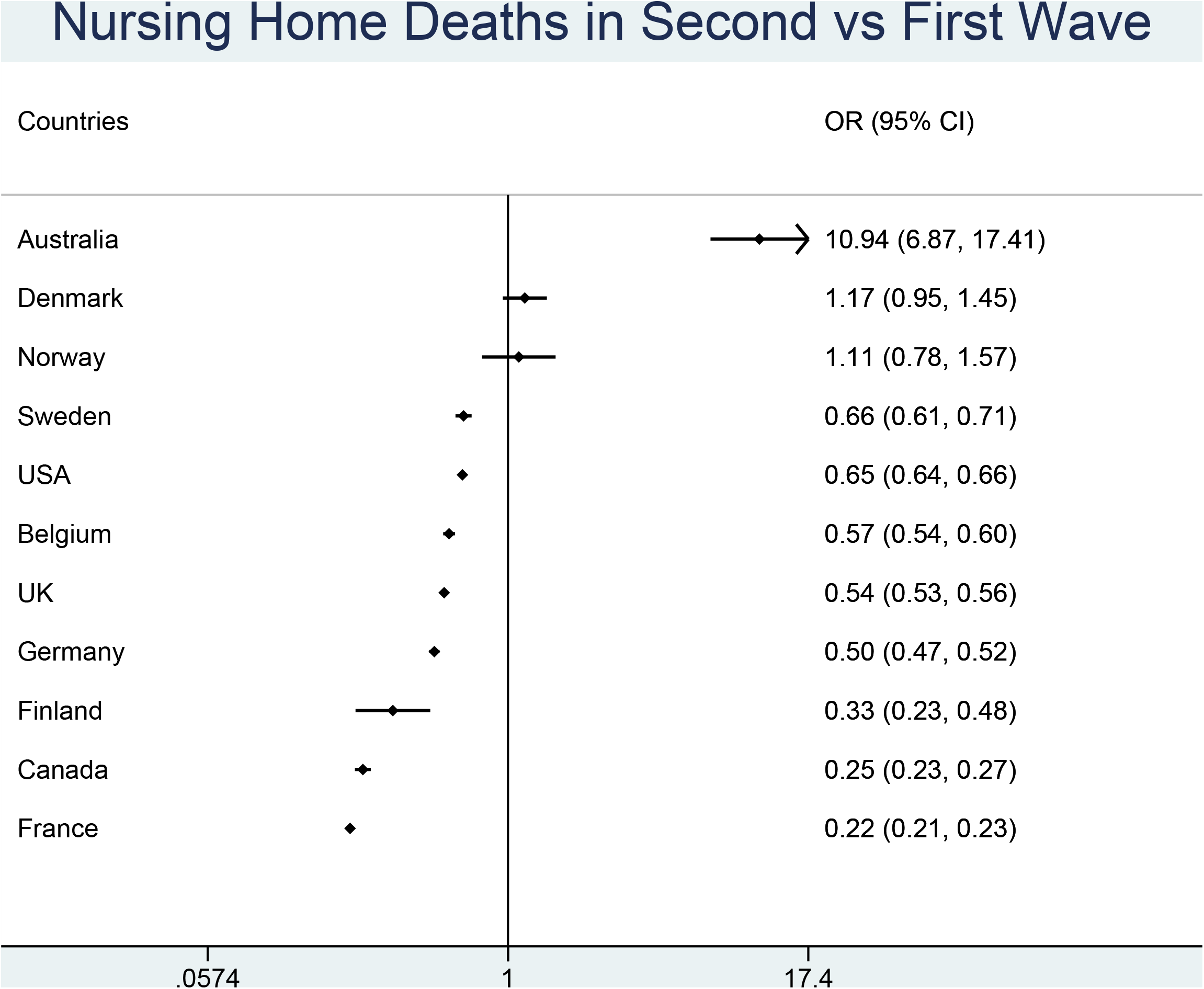

## REFERENCES

1. Williamson E, Walker AJ, Bhaskaran KJ, Bacon S, Bates C, Morton CE, et al. OpenSAFELY: factors associated with COVID-19-related hospital death in the linked electronic health records of 17 million adult NHS patients. medRxiv. 2020:2020.05.06.20092999.

2. Ioannidis JPA, Axfors C, Contopoulos-Ioannidis DG. Population-level COVID-19 mortality risk for non-elderly individuals overall and for non-elderly individuals without underlying diseases in pandemic epicenters. Environmental research. 2020;188:109890.

3. O’Driscoll M, Dos Santos GR, Wang L, Cummings DAT, Azman AS, Paireau J, et al. Age-specific mortality and immunity patterns of SARS-CoV-2. Nature. 2020.

4. Ioannidis JPA. Precision shielding for COVID-19: metrics of assessment and feasibility of deployment. medRxiv. 2020:2020.11.01.20224147.

5. Gudbjartsson DF, Helgason A, Jonsson H, Magnusson OT, Melsted P, Norddahl GL, et al. Spread of SARS-CoV-2 in the Icelandic Population. The New England journal of medicine. 2020.

6. Comas-Herrera A, Zalakaín J, Lemmon E, Henderson D, Litwin C, Hsu A, et al. Mortality associated with COVID-19 in care homes: international evidence. Article in LTCcovid.org, International Long-Term Care Policy Network, CPEC-LSE, 14 October2020.

7. Smith GD, Spiegelhalter D. Shielding from covid-19 should be stratified by risk. BMJ. 2020;369:m2063.

8. Worldometers.info. COVID-19 CORONAVIRUS PANDEMIC Dover, Delaware, USA2020 [updated November 26, 2020. Available from: https://www.worldometers.info/coronavirus/#countries.

9. Comas-Herrera A, Zalakaín J, Litwin C, Hsu A, Lemmon E, Henderson D, et al. Mortality associated with COVID-19 in care homes: early international evidence. Article in LTCcovid.org, International Long-Term Care Policy Network, CPEC-LSE, 26 June 2020.

10. StataCorp. Stata Statistical Software: Release 15. College Station, TX: StataCorp LLC; 2017.

11. Burton JK, Bayne G, Evans C, Garbe F, Gorman D, Honhold N, et al. Evolution and effects of COVID-19 outbreaks in care homes: a population analysis in 189 care homes in one geographical region of the UK. The Lancet Healthy Longevity. 2020;1(1):e21–e31.

12. Salcher-Konrad M, Jhass A, Naci H, Tan M, El-Tawil Y, Comas-Herrera A. COVID-19 related mortality and spread of disease in long-term care: a living systematic review of emerging evidence. medRxiv. 2020:2020.06.09.20125237.

13. Horby P, Lim WS, Emberson JR, Mafham M, Bell JL, Linsell L, et al. Dexamethasone in Hospitalized Patients with Covid-19 - Preliminary Report. The New England journal of medicine. 2020.

14. Axfors C, Schmitt AM, Janiaud P, van ‘t Hooft J, Abd-Elsalam S, Abdo EF, et al. Mortality outcomes with hydroxychloroquine and chloroquine in COVID-19: an international collaborative meta-analysis of randomized trials. medRxiv. 2020:2020.09.16.20194571.

15. The Atlantic Monthly Group. The COVID Tracking Project at The Atlantic: The Long-Term Care COVID Tracker. https://covidtrackingcom/data/long-term-care (Accessed 2021-01-21).

16. Cousins S. Experts criticise Australia’s aged care failings over COVID-19. The Lancet. 2020;396(10259):1322–3.

17. Liu M, Maxwell CJ, Armstrong P, Schwandt M, Moser A, McGregor MJ, et al. COVID-19 in long-term care homes in Ontario and British Columbia. Canadian Medical Association Journal. 2020;192(47):E1540.

18. Spiegelhalter D. Use of “normal” risk to improve understanding of dangers of covid-19. BMJ. 2020;370:m3259.

19. Ministry of Health. COVID-19 Weekly Situation Report 29/06/2020 - 05/02/2020 Turkey. https://covid19.saglik.gov.tr/; 2020.

20. Ioannidis JPA. Global perspective of COVID-19 epidemiology for a full-cycle pandemic. European journal of clinical investigation. 2020;50(12):e13423.

21. Instituto Superiore di Sanità (ISS). Caratteristiche dei pazienti deceduti positivi all’infezione da SARS-CoV-2 in Italia. Dati al 11 november 2020. https://www.epicentro.iss.it/en/coronavirus/sars-cov-2-integrated-surveillance-data; 2020.

22. Santé publique France. COVID-19. Point épidémiologique hebdomadaire du 12 novembre 2020. https://www.santepubliquefrance.fr; 2020.

23. Rijksinstituut voor Volksgezondheid en Milieu – RIVM. Epidemiologische situatie COVID-19 in Nederland, 10 november 2020, 10:00. https://www.rivm.nl; 2020.

24. Instituto Nacional de Salud. COVID-19 en Colombia. http://www.ins.gov.co/Noticias/Paginas/coronavirus-casos.aspx Accessed 2021-01-24.

25. Gobierno Digital Ministerio Secretaría General de la Presidencia, Ministerio del Interior y Ministerio de Ciencia Tecnología Conocimiento e Innovación. Cifras Oficiales COVID-19 en Chile: La Realidad Nacional en Datos. https://www.gob.cl/coronavirus/cifrasoficiales/ Accessed 2021-01-24.

26. Komite Penanganan COVID-19 Dan Pemulihan Ekonomi Nasional. Situasi virus COVID-19 di Indonesia. Peta Sebaran. https://covid19.go.id/peta-sebaran Accessed 2021-01-24.

